# Analysis of Postmortem Dermal Extracts is an Efficient Illicit Drug Screening Method Supporting Forensic Medicine

**DOI:** 10.1101/2025.07.16.25331634

**Authors:** Daniel Wasinger, Katharina Stolz, Michael Wolf, Günter Gmeiner, Andrea Bileck, Samuel M. Meier-Menches, Fabian Kanz, Christopher Gerner

## Abstract

The forensic screening of illicit drugs usually depends on analyzing tissue homogenates, which is resource-intensive. Exploring a non-invasive sampling method using dermal extracts collected postmortem, we developed a rapid technique to analyze these extracts using mass spectrometry coupled with liquid chromatography. Samples from 93 deceased individuals were analyzed alongside a standard tissue-based toxicological assay, which resulted in a total of 312 positive identification events comprising 19 different drugs and drug metabolites commonly associated with illicit use. Referring to results of the tissue-based toxicology assay as reference, the average positive overlap of drug identification in dermal extracts was 95%, and the average negative overlap was 87%, indicating remarkable sensitivity and specificity. While cocaine was a frequent contaminant of dermal extracts, cocaine consumption was specifically assessed by including endogenous cocaine metabolites consistently found across multiple sampling sites. Furthermore, detecting ethyl glucuronide evidenced alcohol consumption, and cocaethylene indicated simultaneous cocaine and alcohol consumption. In 8 individuals, substantial evidence of cocaine consumption was thus collected, despite negative results from the toxicological assay. Notably, patterns of mixture intoxications belonging to drug abuse were separated from natural deaths with a simple scoring system based on illicit drugs and metabolites, demonstrating the support of forensic assessment with dermal extracts. Furthermore, an additional 64 medications and related metabolites were identified in the dermal extracts, resulting in a total of 82 compounds aligning with the toxicological screening. Together, this study demonstrated that the collection and analysis of dermal extracts offers a non-invasive and highly efficient method for postmortem forensic investigation independent of a standard autopsy.

**Highlights:**

- Non-invasive postmortem drug screening method
- Dermal extracts enable economical and flexible sample collection
- Broad coverage of medications, illicit drugs and drug metabolites
- Assessment of combination intoxications

## Introduction

Forensic toxicology, *i.e.* the application of toxicology in forensic investigations, is a crucial component in postmortem examination. It involves the analysis of biological samples to measure the presence of drugs, alcohol, poisons and other toxic substances and to interpret their possible effects on an individual, including their contribution to death. The establishment of novel methods in postmortem examinations, however, is strongly influenced by their practical implementation [1]. Typically, validated tests for verifying illicit drug intoxication require time-consuming and elaborate sample collection and preparation methods [2]. This is true for most gold standard methods, which are mainly based on gas chromatography-mass spectrometric (GC-MS) analysis of tissue extracts [3], but also for the latest generation of immunoassays [4]. Since postmortem examinations legally require trained personnel, specialized equipment and expert documentation, the costs of determining the presence of illicit drugs are also impacted by the laborious methods of sample collection [5]. Despite the increasing recognition of forensic toxicology in postmortem examinations, its widespread use is therefore limited by economic considerations [6]. Therefore, non-invasive sample collection procedures to generate reliable information without the necessity of a resource-intensive autopsy are of great interest [7]. Hair analysis represents a viable source for detecting drugs of abuse in individuals in a non-invasive manner [8–10]. Drug intoxication can be verified over extended exposure windows by analyzing hair using immunoassays or liquid chromatography mass spectrometry (LC-MS)-based methods [11]. Interestingly, the deposition of drugs of abuse in hair is believed to be driven by their incorporation during formation of the hair shaft *via* diffusion from sweat [8]. This process motivated the investigation of sample collection directly from dermal extracts during postmortem examinations because small molecules in eccrine sweat are also thought to be deposited on the skin. Specifically, metabolic analysis of dermal extracts is believed to provide a more acute snapshot of the exposure to illicit drugs compared to hair. We have previously described that the minute amounts of sweat collected from fingertips contains sufficient analyte quantities to detect numerous xenobiotics and endogenous metabolites enabling human biomonitoring [12]. The identification of therapeutic drugs in sweat samples from the fingertip and of drugs of abuse from a fingerprint has also been demonstrated [13–15].

To this aim, a study was designed to collect and analyze dermal extracts from deceased individuals that had undergone the standard procedure of clinical postmortem examination. Dermal extracts were collected from both the palms and soles and were subsequently analyzed by an adapted LC-MS method, similar to that previously used for finger sweat analysis [12]. Here, illicit and therapeutic drugs and their metabolites were screened using an untargeted metabolomics approach as a discovery tool and, where possible, verified against chemical standards. This approach allowed us to identify exposure to illicit drugs *via* dermal extracts. These findings were compared to a validated toxicology assay to evaluate the method. Dermal extracts offer practical advantages as a sample collection method and the mass spectrometry-based analysis may provide additional data when compared to classical toxicological assessments. Thus, this approach might become a useful initial screening method for intoxication during postmortem examinations.

## Material and Methods

### Chemicals and Materials

LC-MS CHROMASOLV methanol (MeOH) (Honeywell, GER), HiPerSolv CHROMANORM water (VWR chemicals, FR), and HiPerSolv CHROMANORM formic acid (FA) (VWR chemicals, UK) was used during sample preparation and LC-MS/MS analysis. As chemical standards, CEDIA® multiple drug calibrators containing benzoylecognine, EDDP, D-methamphetamine, morphine, nitrazepam, phencyclidine and secobarbital were used (Thermo Scientific, Austria).

Sampling units were punched from precision wipes (11 × 21 cm, Kimtech Science, Kimberly-Clark Professional, USA) using a steel puncher of 1.27 cm diameter. The filter paper was pre-wetted with 3 µl water and stored in 0.5 mL Safe-Lock Tubes (Eppendorf Austria GmbH, AT).

### Cohort description

This study was approved by the ethics committee of the Medical University of Vienna, EK-Nr: 2114/2021, entitled: “Feasibility of sweat sampling in the context of a metabolomics approach in deceased individuals that undergo a judicial autopsy”. Ninety-three individuals undergoing medicolegal autopsy at the Center for Forensic Medicine of the Medical University of Vienna were selected for sampling postmortem dermal extract over a time period of one year. The resulting cohort consisted of 37 female and 56 male individuals of high diversity, ranging from 17 to 94 years of age (median = 55) and displaying BMIs ranging from 15.2 to 46.2 (median = 24.4). In 35 cases (38%) the exact time of death could not be determined (Supplementary Table S1). In cases with known exact time of death, autopsies and sampling was performed on average 8.7 days after death, but no later than 18 days after death. General causes of death were defined by medical examiners as either natural (n = 43), traumatic (n = 23) or through intoxication (n = 27), which were further stratified by more detailed specific causes of death (Supplementary Table S1). Fifty-eight individuals (62%) received emergency medical aid shortly before passing.

### Collection of dermal extracts

All deceased individuals were sampled during the course of the performed autopsy. For establishing feasibility, the first 23 individuals were sampled on the left hand, right hand and right foot for a total of 3 samples per individual. All subsequent individuals were only sampled twice, preferably on the right hand and right foot. In two cases, taking samples from the feet was not possible due to the condition of the body. Dermal extract sample nomenclature patterns consist of the number of the individual and the number of the sampling site on the respective individual (e.g. “69_1” for sample 1 of individual #69, Supplementary Table S1).

For cases 1 – 23, the right hand was rinsed with water for 2 – 3 seconds and waited for air drying to take place. Then, the precision wipe sampling implement was taken out of the Safe-Lock tube and placed onto the thumb before applying 100 µL LC-MS grade water. The index finger was moved into position to pinch the paper substrate in place and some force applied for complete contact. The fingers were held in place by cable binders for 5 minutes before returning the sampling paper to the tube. The left hand was similarly sampled with omission of the washing step at the start. In cases 24 – 94, only one hand and one foot in unwashed state were sampled from each individual. Case 37 was not cleared for toxicological screening and had to be excluded from analysis.

A high-throughput dilute-and-shoot approach was carried out for metabolite extraction from the sample units. Addition of 120 µL extraction solution consisting of water with 0.2% FA was followed by 30 seconds in an ultrasonic bath for homogenously diluting the sweat residue on the sample units. The solution was transferred into HPLC vials equipped with a 200 µL V-shape glass insert (both Macherey-Nagel GmbH & Co.KG, GER) and analysed by LC-MS/MS.

### LC-MS/MS analysis of dermal extracts

A Vanquish UHPLC System (Thermo Fisher Scientific) coupled to a Q Exactive HF (Thermo Fisher Scientific) mass spectrometer was employed for this study. Chromatography was performed using a Kinetex XB-C18 column (100 Å, 100 × 2.1 mm, 2.6 µm, Phenomenex Inc.). Mobile phase A consisted of water with 0.2% formic acid, mobile phase B of methanol with 0.2% formic acid and the following gradient program was run: 1–5% B in 0.3 min, 5–40% B from 0.3–4.5 min, followed by a column washing phase of 1.4 min at 80% B and a re-equilibration phase of 1.6 min at 1% B resulting in a total runtime of 7.5 min. Flow rate was set to 500 µL min^−1^, the column temperature to 40 °C, and the injection volume was 10 µL.

All samples were analysed in technical duplicates. An untargeted mass spectrometric approach was applied. Electrospray ionization was performed in positive and negative ionization mode at 3200 and 2500 V, respectively. Capillary temperature was set to 350 °C, aux gas heater to 400 °C, sheath gas to 60 AU, aux gas to 20 AU, sweep gas to 2 AU (5 AU in –ESI), and S-lens RF level to 30%. MS1 scan range was set to m/z 100–1000, resolution to 60000 FWHM (at m/z 200), AGC-target to 1e6, and maximum injection time to 50 ms. A top 4 ddMS2 approach applying 30% normalized collision energy (NCE) was used for untargeted fragmentation. MS2 scan resolution was set to 15000 FWHM (at m/z 200), isolation window to 1.2 m/z, AGC-target to 1e5, maximum injection time to 50 ms, and dynamic exclusion to 6 s. The instrument was controlled using Xcalibur software (Thermo Fisher Scientific).

### Data Evaluation

Thermo raw files of mass spectrometric data of dermal extracts were analyzed using MacCoss Skyline software (ver. 24.1.0.199). Data was also converted to mzML format using MSConvert (ver. 3.0.22354) from the ProteoWizard Tool Kit. These mzML files were used for the analysis of MS2 spectral data for identification purposes using SIRIUS (ver. 5.8.6) [16]. Finally, all statistical work was done in R Studio using R ver. 4.3.2.

Data received from the toxicological assays about individuals following postmortem medical examination and detected substances was converted into true/false data tables. Using chemical formulas of the detected substances, an investigative mass list was created. This list was employed to search for candidate peaks in the mass spectrometric data of dermal extract samples of individuals with positive reports of the respective substances. Upon confirming viable candidates using the independent MS2-based *de-novo* annotation capabilities of SIRIUS, retention times were documented (Supplementary Table S2). With this information, manual integration was applied to all matching peaks in the remaining dermal extract samples. Results from both the toxicological assay and the dermal extract analysis were finally collated and reorganized into a pivoted format (Supplementary Table S3).

### Sample collection for toxicological assessment

In the course of the autopsy performed by medical examiners, samples for toxicological analysis were taken in accordance with routinely used standard operating procedures. Peripheral blood was obtained from the femoral artery and stored in plastic tubes. Sinus blood was obtained from venous blood vessels from the dissected brain and stored in plastic tubes. Wherever sufficient state of tissue preservation allowed for it, around 30 g of the cerebellum and 2 g of the medulla oblongata were taken and stored separately in plastic containers. In cases where autolytic degradation was too advanced to discern brain regions, 30 g of the homogenized brain mass was taken and stored in a plastic container instead. Following dissection of the stomach, 120 mL of stomach content was taken and stored in plastic tubes. In cases of insufficient volume, the entire stomach content was transferred. Urine was similarly taken in its entirety directly from the dissected urinary bladder. All samples were stored at −22 °C until collection and transport to analytical institutions, except for blood sample aliquots used for ethanol determination, which were stored at +5 °C.

## Results

### Metabolomic analysis of postmortem dermal extracts

The original sample collection method [12] was adapted due to the absence of active sweating in deceased individuals. Specifically, the precision wipes used for collecting dermal extracts were soaked with an additional 100 µL water following their application to the skin and then firmly pressed for 5 min at the indicated location on fingers and soles (Figure 1). The metabolites in the wet sampling unit were then extracted with 120 µL aqueous solution and analyzed by an untargeted metabolomics workflow using a ultra-high performance liquid chromatography-mass spectrometry (UHPLC-MS) method. This efficient workflow can be accomplished within 30 min from sampling to the analysis results, potentially allowing for high throughput (Figure 1). Sample collection from soles turned out to be more practical compared to collection from fingers.

**Figure 1:**
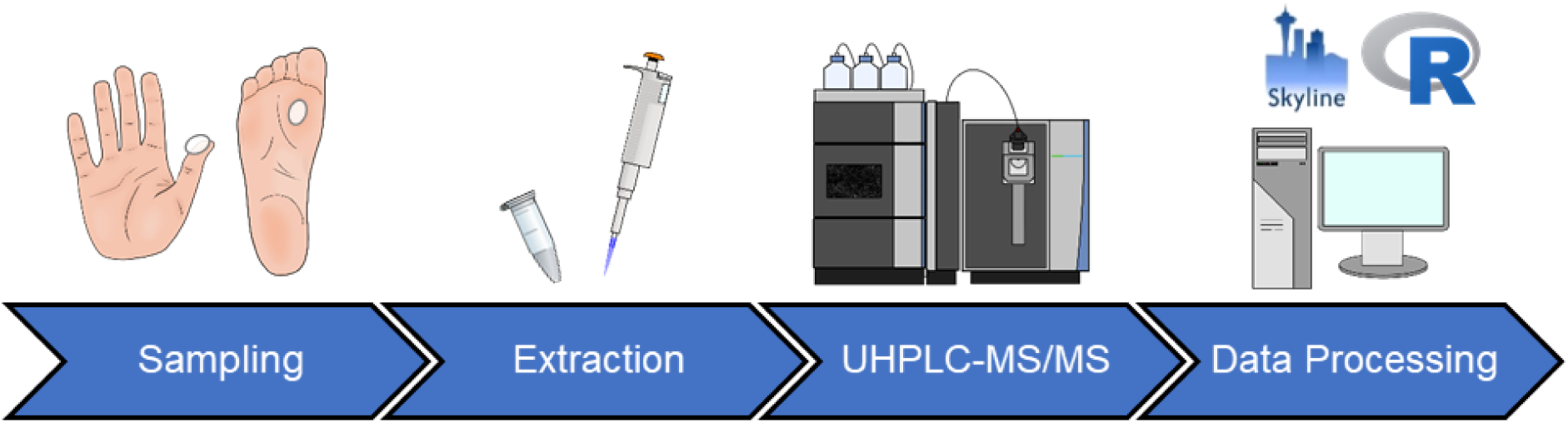
Scheme of the analytical workflow. The entire procedure from sample collection to data processing can be accomplished within half an hour.

Out of the 123 illicit and therapeutic drugs, as well as metabolites that were covered in the toxicological assay, 82 (67%) were identified in dermal extracts, including the most common illicit or controlled substances with abuse potential, e.g. cocaine, codeine, morphine, methadone and fentanyl (Supplementary Table S2). Due to stringent regulations, nine illicit drugs were obtained as certified standard material, while the remaining annotations in dermal extracts remained level 2 identifications.

### Comparison of analysis results with validated toxicological assay

The dermal extracts from the cohort of 93 deceased individuals were analyzed in-depth in relation to 19 substances commonly associated with illicit drug use. These drugs were identified in dermal extracts by key LC-MS-based parameters, including chromatographic retention time, accurate molecular mass and molecular fragment (MS2) spectra, matching the analytical standard spectra (Figure 2, Supplementary Table S2). In cases where standard material was lacking, annotation was further based on chemical fingerprint analysis *via* SIRIUS [16]. Drugs were readily detected in the extracted ion chromatograms (EICs) with defined chromatographic peaks. The presence and absence of specific drugs in individuals was unequivocally determined by the EICs (Figure 2B). Most positive identifications obtained with this method agreed with the toxicological reports except for fentanyl and norfentanyl, and most drugs and metabolites showed 100% positive overlap across the different samples (Table 1). However, sometimes the dermal extracts also identified substances not indicated by the toxicological report, resulting in negative overlap values below 100%. Whether these cases constituted false positives through skin contamination required further investigation.

**Figure 2:**
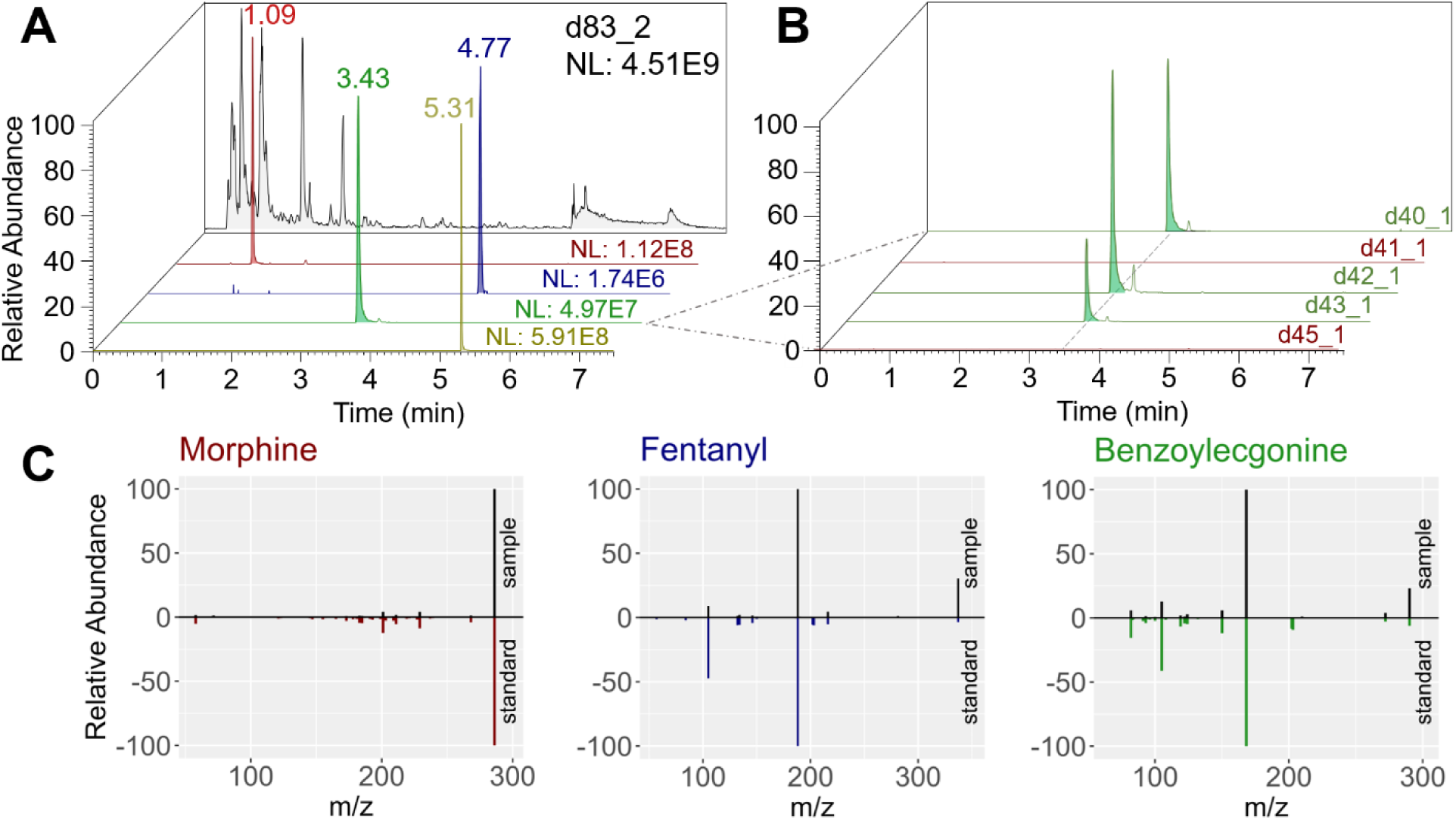
(A) Total ion chromatogram (TIC) and extracted ion chromatograms (EICs) of morphine (red), fentanyl (blue), benzoylecgonine (green), methadone (yellow) and their respective signal intensities (NL) exemplified by sample d83_2. (B) EICs of benzoylecgonine in different samples, showcasing a clear absence of peaks in negative samples (red). (C) Contrasted MS2 spectra of select molecules in samples (above, black) and the corresponding analytical standards (below, colored).

**Table 1:** Computed positive identification overlap and negative identification overlap of drugs and drug metabolites detected in samples of dermal extracts in reference to the toxicological assay (N = 93), including their reported frequency (N°(Tox. reports)) and the frequency found in dermal extracts (N°(Dermal extract). EDDP = 2-Ethylidin-1,5-dimethyl-3,3-diphenylpyrrolidine; MDA = 3,4-methylenedioxyamphetamine; MDMA = methylenedioxymethylamphetamine.

| Molecule | N°(Tox. reports) | N°(Dermal extract) | Positive overlap | Negative overlap |
| --- | --- | --- | --- | --- |
| 6-Acetylmorphine | 2 | 20 | 100.0% | 80.2% |
| Amphetamine | 4 | 19 | 100.0% | 83.1% |
| Benzoyllecgonine | 9 | 37 | 100.0% | 66.7% |
| Cocaethylene | 1 | 16 | 100.0% | 83.7% |
| Cocaine | 6 | 51 | 100.0% | 48.3% |
| Codeine | 7 | 15 | 100.0% | 90.7% |
| Dihydrocodeine | 1 | 6 | 100.0% | 94.6% |
| Ecgonine methyl ester | 8 | 20 | 100.0% | 85.9% |
| EDDP | 2 | 3 | 100.0% | 98.9% |
| Fentanyl | 13 | 13 | 69.2% | 95.0% |
| Ketamine | 12 | 28 | 91.7% | 79.0% |
| MDA | 2 | 4 | 100.0% | 97.8% |
| MDMA | 2 | 5 | 100.0% | 96.7% |
| Methadone | 2 | 3 | 100.0% | 98.9% |
| Methamphetamine | 3 | 16 | 100.0% | 85.6% |
| Morphine | 23 | 30 | 91.3% | 87.1% |
| Norfentanyl | 4 | 5 | 50.0% | 96.6% |
| Normorphine | 8 | 9 | 100.0% | 98.8% |
| O-Desmethyltramadol | 3 | 12 | 100.0% | 90.0% |

### Drug metabolites support the management of surface contamination

Surface contamination may generally represent a potential source for identifying illicit drugs following non-invasive sample collection from hair and skin [8]. Especially cocaine and benzoylecgonine were identified much more frequently in dermal extract than by the toxicological assay. This is unsurprising given that cocaine can be found ubiquitously on public surfaces connected to day-to-day interactions such as public transport, banknotes, coins and door handles [17]. Although benzoylecgonine is the preferred analyte to indicate cocaine consumption in tissue-based assays, the associated danger of skin contamination by touch has been shown to make both cocaine and benzylecgonine poor markers for consumption using fingerprints prior to hand washing [18].

Furthermore, the activities of metabolic enzymes may differ between individuals, which can affect metabolite patterns. Thus, monitoring a larger part of the metabolic network, increases the overall confidence of a positive identification. Leveraging the capabilities of the untargeted analysis method employed for dermal extracts, the analysis was expanded to incorporate additional metabolites of cocaine, *i.e.* ecgonine, hydroxybenzoylecgonine, norbenzoylecgonine and norcocaine (Figure 3). Reinvestigating the dermal extracts of the 9 cases in whom cocaine consumption was indicated by the toxicological assay, 4 or more of the additional metabolites could be confirmed in every individual. Following this, all other cocaine-positive dermal extracts were re-evaluated with regard to these cocaine metabolites. This strategy identified 8 additional cases exhibiting an network of 4 or more cocaine metabolites, resulting in a total of 17 suspected cocaine consumers. Interestingly, all of the additional individuals were also victims of other drug intoxications or polytrauma, featuring multiple different illicit substances or alcohol in their toxicological report. Given the tendency of users of illicit drugs to consume a mixture of substances, this lends further plausibility to the obtained results [19].

**Figure 3:**
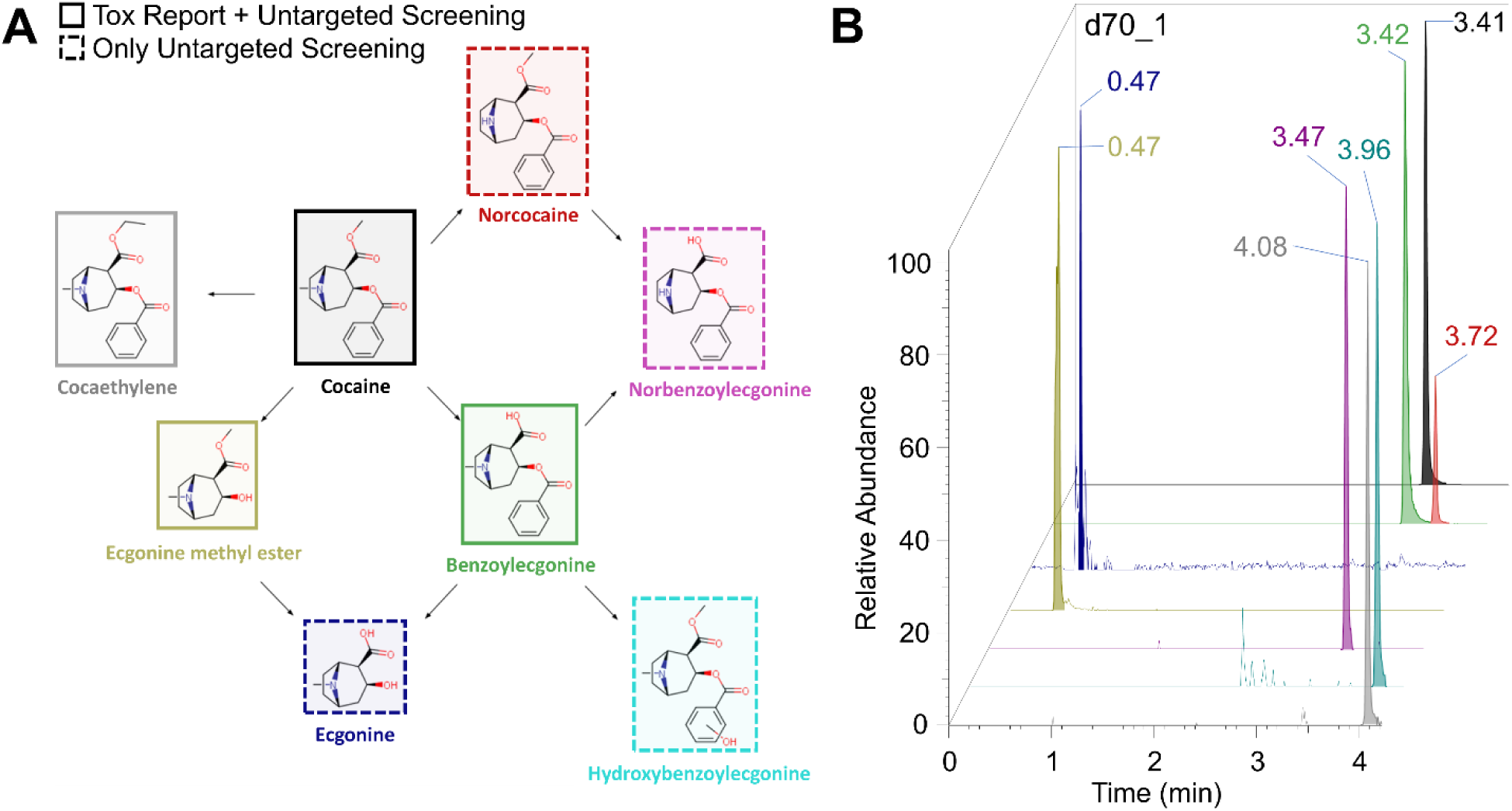
(A) Cocaine and its metabolic breakdown products. Molecules with a solid border denote compounds accessible through both the targeted toxicological assay and the untargeted analysis of dermal extracts. Dashed borders denote compounds only tracked in the dermal extracts. (B) The extracted ion chromatograms show color-matched signals respective to described compounds in A in sample d70_1 as an example.

Cocaethylene is a conditional metabolite of cocaine that forms through substitution of the methane moiety at the ecgonine carboxylic group [20]. While ethanol and its major metabolites cannot be directly determined with the established method, typically under 0.1% of ethanol is directly conjugated with glucuronic acid within the body, forming ethyl glucuronide [21], which we found to be detectable in dermal extracts and thus indicated alcohol consumption. Out of the 17 suspected cocaine consumers, 12 were reported to have ethanol in blood or urine by the toxicological analysis. In 11 of these cases, both ethyl glucuronide and cocaethylene were identified in dermal extracts. Remarkably, these signals were stable down to the lowest reported ethanol concentration of 0.01 per mille.

Since at least two dermal extract samples were collected for each individual, metabolites were also evaluated on their detection consistency. Ecgonine methyl ester, also available in the toxicological screening, was the most consistent metabolite, exhibiting clear signals across all dermal extract samples of the 17 individuals. Hydroxybenzoylecgonine and norcocaine also proved to be robustly detectable in all matched samples across 15 of those 17 individuals, while ecgonine and norbenzoylecgonine were less reliable and were not detectable in 3 and 2 cases, respectively.

Overall, this data demonstrated that dermal extracts detected cocaine consumption in cases not covered by the toxicological assay and that the analysis of drug metabolites successfully addresses the challenge of surface contamination, which could be further mitigated by sample collection at multiple locations on the skin.

### Dermal extract drug profiles relate to specific causes of death by intoxication

An important part of forensic analysis is to provide evidence, if an individual had consumed illicit substances or not. Using standard procedures and the results from the toxicological assays, the 93 deceased individuals in the present study were classified according to the cause of death, distinguishing trauma, natural death, and intoxication (Supplementary Table S1). Using dermal extracts, positive identifications of the substances listed in Table 1 were summed up to create a drug score for each individual. As discussed previously, for lack of robustness cocaine and benzoylecgonine were removed and replaced with metabolites from the extended cocaine metabolite network. Furthermore, to avoid score inflation of drugs featuring multiple breakdown products, the results were scaled by dividing molecule identifications belonging to the same precursor drug by the amount of metabolites tracked for a maximum contribution of 1. As morphine is both a drug but also a breakdown product of codeine and 6-acetylmorphine, this method slightly undervalues contributions coming from morphine consumption but still gives a good estimation of drug use per individual. Figure 4A shows histograms of calculated drug scores for individuals separated by the general cause of death with the average score indicated by vertical dashed lines. Expectedly, the average drug score of intoxication deaths is higher than those of natural or trauma deaths. However, there are also cases of intoxication with very low and zero scores. It may be noted that the intoxication class also included non-drug-related intoxications, *e.g.* carbon monoxide poisoning and medical drug intoxication. Figure 4B shows an additional differentiation of intoxication deaths between drug-related and other causes, further separating the groups. Natural deaths are most frequent on the low score edge with some instances of higher scores. Many commonly abused drugs have accepted medicinal use, especially for pain control, making the appearance of single substances not unlikely. Furthermore, given the widespread societal use of illicit drugs, it is reasonable to find functional, recreational users that happened to be under the influence at the time of death while dying due to other causes. This pattern is reflected in the deaths by trauma, showing examples of all previously discussed cases.

**Figure 4:**
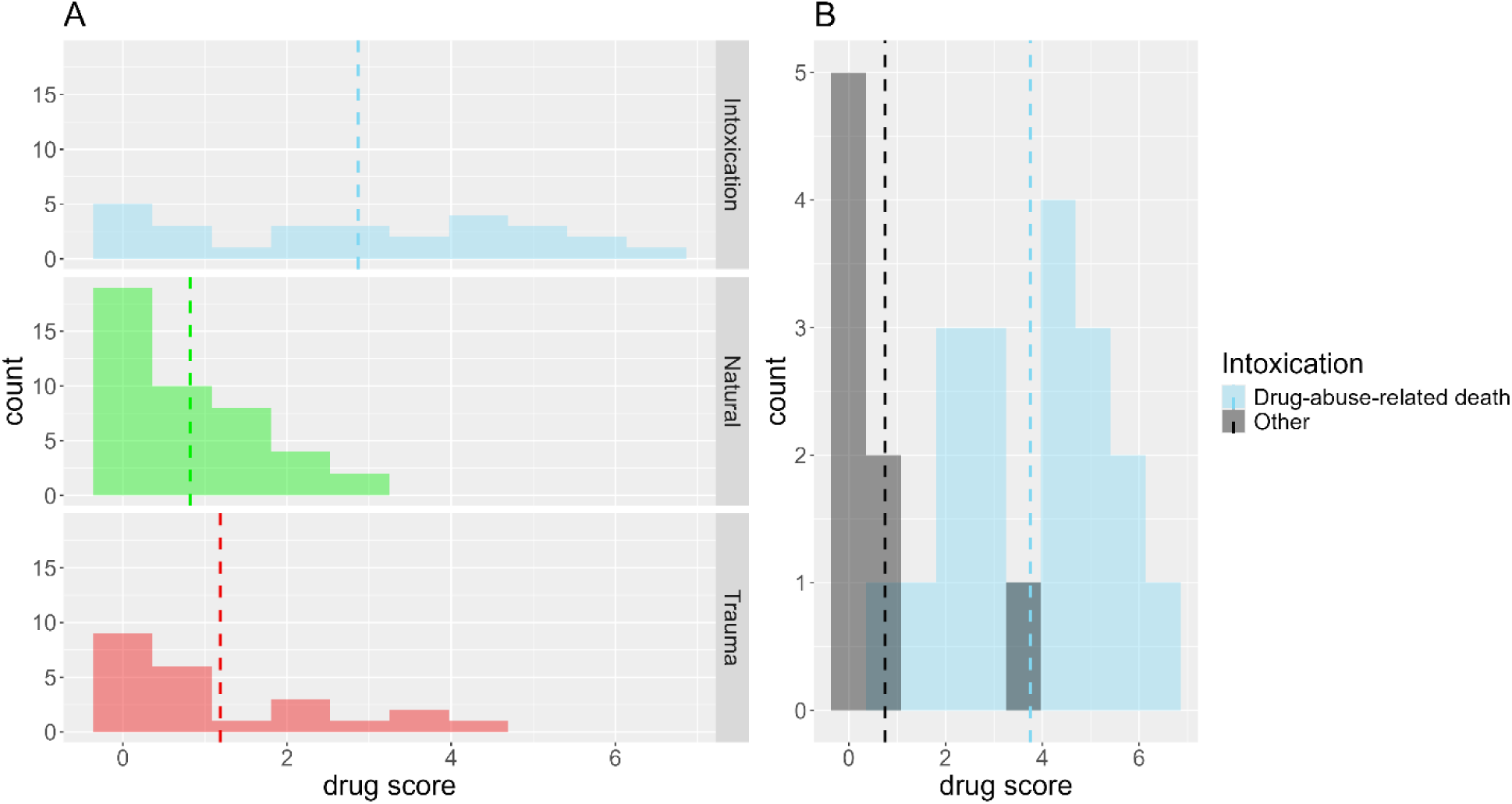
(A) Histograms of calculated drug scores, separated by general causes of death. Vertical dashed lines indicated average scores. (B) Histogram of calculated drug scores for deaths by intoxication, further stratified by drug-abuse-related deaths and other intoxications, such as carbon monoxide poisoning or medical drug intoxication.

To further investigate the individual profiles of illicit and therapeutic drugs, all drugs detected by dermal extracts were also classified into functionally related groups of compounds (Supplementary Table S2) and their frequency in relation to specific causes of death was computed. In detail, the unspecific mixture intoxication group was dominated by cocaine metabolites, opioids, benzodiazepines and CNS stimulants, with a sizeable contribution from antidepressants, antipsychotics and alcohol markers (Figure 5). Notably, the groups cocaine intoxication, methamphetamine intoxication and morphine intoxication showed a very similar pattern to mixture intoxication. Again, this may be expected as it is known that users of illicit drugs tend to consume mixtures of substances [19].

**Figure 5:**
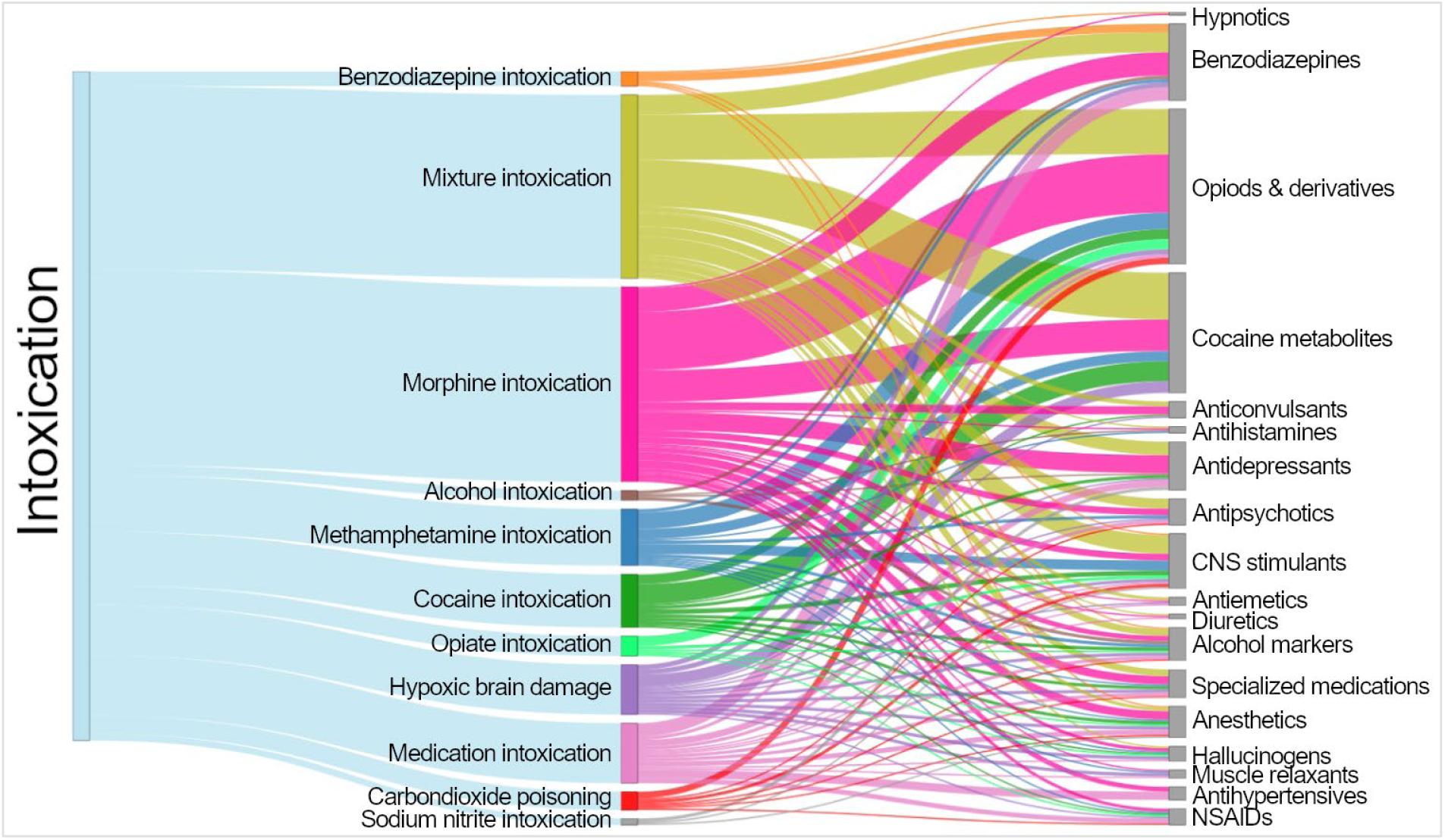
Alluvial plot connecting groups of intoxication and the respective classes of drugs positively detected in the forensic dermal extracts sample set according to relative frequency.

In principle, dermal extracts may thus detect frequent mixture intoxications in a rapid manner, providing valuable evidence to examiners for the need of a validated forensic assay.

## Discussion

We present a simple sample collection and analysis method for identifying illicit and therapeutic drugs, as well as their metabolites, from postmortem dermal extracts. Reliable and detailed information is crucial for making informed decisions in forensic contexts. However, traditional sample preparation methods used in classical toxicology can hinder the wider application of drug screening due to considerable costs. Collection of dermal extracts for postmortem metabolic analysis presented here does not require an autopsy and may help in overcoming this limitation. Indeed, a total of 82 small molecules, related to illicit and therapeutic drugs and their metabolites, were identified by collecting dermal extracts of 93 deceased individuals, underscoring its potential use for forensic toxicology.

To determine the reliability of the assay, we compared the results from dermal extracts to those from classical tissue-based toxicology in regard to commonly abused illicit and controlled drugs. Dermal extracts achieved excellent overlap with the information provided by the toxicological assay, with the exception of fentanyl and norfentanyl. Fentanyl is known to be 50 to 100 times more potent than morphine and 2 mg can be lethal for the average adult [22]. As such, quantities are expected to be much lower than other drugs. In any case, untargeted mass spectrometry-based analysis is useful for screening and discovery purposes, but not for validated methods [23]. Establishing targeted analytical techniques, tailored to specific forensic needs, will likely also further expand the range of detectable drugs and their metabolites in dermal extracts and lower the limit of detection for low-abundant substances such as fentanyl.

Notably, the dermal extracts provided reliable evidence of cocaine consumption when the validated assay did not. We have previously described dynamic profiles of anticancer drugs in sweat of the fingertips of patients receiving chemotherapy [14]. The identified drug cyclophosphamide was detectable for prolonged times following administration, exceeding the reported half-lifes in plasma. These findings may suggest that drugs are deposited in the epidermis in a manner similar to hair [24], but the epidermis probably reflects a more acute depot of drug exposure. This suggests, that dermal extracts could be more sensitive in cases of continued or habitual consumption. Indeed, this could explain the lower negative overlap on the basis of the toxicological assay data. Of course, the non-invasive sample collection from hair or dermal extracts is potentially prone to surface contamination [25]. It was already suggested that surface contamination could be mitigated by also considering endogenous metabolites [26]. From a total of 51 cases with positive cocaine identification in dermal extracts, we identified 17 cases in which an extensive network of at least 4 or more cocaine metabolites was detected. These 17 cases also included the 9 positively identified by the toxicological assay. Ecgonine methyl ester, hydroxybenzoylecgonine and norcocaine appear to be the most reliable cocaine metabolites in dermal extracts, with very consistent detection regardless of sampling site. The simultaneous detection of cocaethyle and ethyl glucuronide, as conditional metabolites of alcohol coexposure, further corroborated the findings. Therefore, surface contamination can be efficiently addressed by including these metabolites and sampling at least at two locations, e.g. on the palms and soles. The final establishment of thresholds discriminating between drug consumption and surface contamination will require more empirical data.

The analysis of dermal extracts may offer significant advantages for forensic assessments. The toxicity of certain drugs can lead to fatal outcomes, particularly in cases of combined drug intoxication, which may result from repeated substance abuse [27]. Indeed, the frequency of detection in dermal extracts of certain drug combinations with fatal outcome may inform about their toxicity and, potentially, contribution to death. Incorporating the purely qualitative dermal extract data on commonly abused illicit and controlled substances as well as metabolites into a weighted drug score allowed for good separation of deaths related to drug abuse and other intoxications. Pointing out the nature of the distributions seen in the causes of death may seem trivial, if not for the fact that they are based solely on dermal extracts. This shows that dermal extracts could act as a basis for providing critical evidence to important forensic questions without the need of laborious and costly autopsies or, at the very least, give grounds to request such autopsies. We are currently exploring whether drugs consumed several days prior to sampling can still be detected in dermal extracts, potentially extending the relevant information extracted for forensic evaluations. In any case, the depot function of the epidermis with respect to drug accumulation apparently enabled the identification of illicit and therapeutic drugs for at least up to 16 days after death, while the average time delay from death to postmortem analysis was 8.3 days (Supplementary Table S1).

Additionally, while the drug profiles relating to specific causes of death were used purely descriptive in the scope of this study, classification strategies on the basis of logistic regression algorithms could be explored [28]. These models would provide likelihood ratios for common scenarios encountered in forensic investigations, such as habitual drug-abuser vs. non-user, and in turn more accurately guide the decision process of forensic personell [29]. This data-driven approach would give crucial contextual information to address problems with cognitive biases, shown to have a significant impact on forensic science [30].

The advantages of this assay are also associated with certain limitations. Not all drugs covered by the toxicological assay were readily detected in the dermal extracts. It is not yet established whether certain drugs may generally fail to become detectable in dermal extracts. Furthermore, the presented study focused entirely on qualitative data obtained from dermal extracts. The quantitative data exhibited substantial variability due to the absence of conventional normalization parameters in the sample collection, such as volume or weight. Nevertheless, analogous to the normalization of urine samples to creatinine, it may be feasible to significantly reduce this variability through similar strategies to be established in the future, thereby enabling the extraction of quantitative information from dermal extracts [31].

## Conclusions

We found that dermal extracts obtained from deceased individuals can be used for non-invasive forensic toxicological analysis without the need for an autopsy. Although samples were collected from the fingertips and soles of the feet, sample collection from soles turned out to be more practical during postmortem examination. The newly developed method effectively detected a wide range of drugs of abuse, therapeutic drugs, and their metabolites that are typically identified through more complex and labor-intensive toxicological assays. Postmortem dermal extracts featured remarkable sensitivity in detecting drugs of abuse compared to the conventional toxicological assay, while the specificity will need to be improved in the future. Inclusion of endogenous metabolites of drugs of abuse aided in ruling out false positive identifications by surface contamination, as did the sample collection at multiple locations, e.g. finger and soles, show by the example of cocaine. While alcohol consumption was detected by ethyl glucuronide, cocaethylene specifically indicated the simultaneous intoxication with cocaine and alcohol. Consequently, this method makes it possible to track comprehensive metabolic pathways of selected drugs directly from deceased individuals.

The unique combination of efficiency and sensitivity makes this method particularly suitable for postmortem drug screening, offering low costs and quick data turnaround. Moreover, it has the potential to be integrated with machine learning and artificial intelligence for enhanced decision-making.

## Data Availability

All data produced in the present study are available upon reasonable request to the authors.

## Acknowledgements

The authors are grateful to the Core Facility of Mass Spectrometry at the Faculty of Chemistry and the Joint Metabolome Facility, both members of the Vienna Life-Science Instruments (VLSI).

## Author contributions

KS and FK collected samples. DW, KS and MW performed sample preparation. DW, MW and GG acquired and processed data. AB, SMM, and CG processed data. FK, SMM and CG planned and supervised the study. DW and CG wrote the initial draft of the manuscript, which was edited and approved by all authors.

## Supplementary Material

Supplementary Table S1: Cohort description and cause of death.

Supplementary Table S2: List of analytes, their m/z values, sum formula, polarity and retention time.

Supplementary Table S3: List of compound identifications in the cohort

